# Severe haemolysis during primaquine radical cure of *Plasmodium vivax* malaria: two systematic reviews and individual patient data descriptive analyses

**DOI:** 10.1101/2023.03.02.23286587

**Authors:** Daniel Yilma, Emily S Groves, Jose Diego Brito-Sousa, Wuelton M Monteiro, Cindy Chu, Kamala Thriemer, Robert J Commons, Marcus V G Lacerda, Ric N Price, Nicholas M Douglas

## Abstract

**Background:** Primaquine (PQ) kills *Plasmodium vivax* hypnozoites but can cause haemolysis in patients with glucose-6-phosphate dehydrogenase (G6PD) deficiency.

**Methods:** We did two systematic reviews: the first used data from clinical trials to determine the spectrum of definitions and frequency of haematological serious adverse events (SAE) related to PQ treatment of vivax malaria. The second used data from prospective studies and case reports to describe the clinical presentation, management and outcome of ‘severe’ PQ-associated haemolysis necessitating hospitalisation.

**Findings:** In the first review, SAEs were reported in 70 of 249 clinical trials. There were 34 haematological SAEs amongst 9,824 patients with vivax malaria treated with PQ, 9 of which necessitated hospitalisation or blood transfusion. Criteria used to define SAEs were diverse. In the second review, 21 of 8,487 articles screened reported 163 patients hospitalised following PQ radical cure; 79.9% (123/154) of whom were prescribed PQ at ≥0.5mg/kg/day. Overall, 101 patients were categorised as having probable or possible ‘severe’ PQ-associated haemolysis, 96.8% of whom were G6PD deficient (<30% activity). The first symptoms of haemolysis were mostly reported on day 2 or 3 (45.5%) and all patients were hospitalised within 7 days of PQ commencement. 57.9% (77/133) of patients had blood transfusion. Seven (6.9%) patients with probable or possible haemolysis died.

**Interpretation:** Even when G6PD testing is available, enhanced monitoring for haemolysis is warranted following PQ treatment. Clinical review within the first 5 days of treatment may facilitate early detection and management of haemolysis. More robust definitions of severe PQ-associated haemolysis are required.

**Funding:** WHO-TDR, Australian National Health and Medical Research (NHMRC), The Bill & Melinda Gates Foundation.

## Introduction

*Plasmodium vivax* forms dormant liver stages (hypnozoites) that can reactivate periodically, causing recurrent blood stage malaria infections known as relapses. Primaquine (PQ) is an 8-aminoquinoline drug that has been used for malaria treatment and prophylaxis since the 1950s. It is the only widely available antimalarial that eliminates *P. vivax* hypnozoites and thus can prevent relapses. The World Health Organization (WHO) recommends PQ for radical cure of *P. vivax* or *P. ovale* malaria in all malaria transmission settings in children and adults except pregnant women, infants aged <6 months, women breastfeeding infants <6 months and women breastfeeding older infants with unknown glucose-6-phosphate dehydrogenase (G6PD) status.^1^

PQ can cause severe drug-induced haemolysis, particularly in individuals predisposed to oxidant erythrocytic damage due to functional deficiency of the enzyme G6PD.^2-4^ G6PD deficiency is the commonest human enzymopathy globally and is particularly prevalent in malaria endemic regions.^5,6^ Patients with PQ-induced haemolysis can develop severe anaemia necessitating blood transfusion and haemoglobinuria, potentially leading to kidney damage.^7^ If severe, PQ-associated haemolysis can be fatal.^8^

The WHO recommends G6PD activity assessment prior to administration of PQ for radical cure,^1^ but there is limited access to practicable assays in malaria endemic regions.^9^ PQ treatment is therefore often not implemented due to the risks of haemolysis with unguided use.^10^ Conversely patients with *P. vivax* malaria who are not treated with PQ radical cure can have major deleterious consequences from multiple relapses, including severe anaemia.^11,12^ Policymakers therefore need to weigh potential risks of PQ-associated haemolysis against the risks of recurrent malaria when deciding on treatment protocols. To do this, they need robust data on the incidence and outcomes of PQ-attributable severe haemolysis.

The diagnosis of haemolysis secondary to PQ is typically based on a fall in haemoglobin and/or clinical manifestations suggestive of intravascular haemolysis. Definitions for ‘severe’ or clinically significant PQ-associated haemolysis are not established. Attribution of haemolysis to PQ is difficult in the presence of concomitant parasitaemia.

We did two complementary systematic reviews to inform and standardise PQ pharmacovigilance practices and to facilitate quantification of the risks and benefits of different radical cure strategies. The aim of the first review was to determine the spectrum of definitions and frequency of haemolytic serious adverse events (SAEs) in patients enrolled into prospective *P. vivax* antimalarial clinical efficacy trials. The second review used individual patient data from case reports and prospective studies to describe the clinical presentation, management and outcome of patients with severe PQ-associated haemolysis who required hospitalisation for clinical management.

## Methods

### Review 1 - Serious adverse events in clinical trials

The first systematic review used data from the WorldWide Antimalarial Resistance Network (WWARN) “vivax surveyor”.^13^ This repository contains details of all *P. vivax* antimalarial clinical efficacy studies done between Jan 1960 and August 23, 2021 (previously registered at PROSPERO [CRD42016053228]). To be eligible for inclusion in the current review, trials had to have been done since 1990 and include one or more treatment arm(s) in which patients were treated with either partial or fully supervised PQ therapy for at least 5 days (Box 1 - appendix pp5). PQ administration had to commence within 7 days of starting blood schizontocidal therapy. Data from non-PQ-containing arms in these studies were also extracted for comparative purposes. Studies meeting the above criteria, but not reporting the presence or absence of adverse effects of treatment, were excluded.

Generic definitions of <u>serious adverse events</u> (SAEs) have been standardised^14^ and generally speaking encompass events that are life-threatening or require hospitalisation. In this review, the respective authors’ classification of events as SAEs was accepted but the clinical and laboratory criteria used by the authors to classify the events as SAEs were outcomes of interest.

Data were extracted into Microsoft Excel^®^ including the number of patients enrolled into each treatment arm, patient demographics, clinical and laboratory features, the number of patients experiencing any author-reported SAE, the relationship of SAE to PQ administration and the number of haematological SAEs. ‘Severe’ haematological adverse events (defined *a priori* as a haematological event requiring hospitalisation, blood transfusion, renal replacement therapy or death) were assigned after data extraction based on the clinical data available. Original investigators were approached for missing data and where possible the clinical and laboratory data used by the original investigators to classify episodes as haematological SAE were collected for individual patients. Laboratory methods used to screen for G6PD deficiency (if performed) and individual G6PD activity measurements of patients experiencing a haematological SAE were also collected.

### Review 2 - Severe primaquine-associated haemolysis

In the second systematic review, all articles in any language reporting at least one case of severe PQ-associated haemolysis were identified. To be eligible for inclusion, papers had to report data attributable to individual patients receiving PQ for *P. vivax* radical cure or terminal prophylaxis. The review was registered at PROSPERO [CRD42020196604]. The term **<u>severe haemolysis</u>** was used to define a clinical event requiring hospitalisation, blood transfusion, renal replacement therapy or death, which the original investigators attributed to haemolysis or anaemia. Children <6 months of age and pregnant or lactating women were excluded from the analysis as both are ineligible for PQ treatment.

We searched PubMed, Web of Science, Embase and the Cochrane Central Database for eligible reports published between 1 January 1940 and 20 May 2020, according to PRISMA guidelines (PRIMSA Checklist - appendix pp2-4). The search terms are presented in (box 2 appendix pp6). We also searched articles included in the WWARN *P. vivax* clinical trial database used in the first systematic review,^13^ as well as reference lists from identified articles and documents, conference abstracts and unpublished works from our network of collaborators. The review process was undertaken by six independent reviewers (DY, EG, KT, RJC, NMD, RNP), with discrepancies resolved by discussion.

Data from relevant reports were extracted into a standardised RedCap^®^ database, including the source of data, patient demographics, PQ dose regimen, G6PD activity and all available clinical and laboratory features of the haemolytic event (see Supplementary Data File). Authors of eligible studies were contacted and asked to provide relevant additional information that was not reported in the publication. All data were de-identified. From the data available, the certainty of the diagnosis of PQ-associated severe haemolysis was categorised as “Probable”, “Possible” or “Uncertain” using an *a priori* hierarchical evidence scale (see box 1).

### Statistical analyses

Statistical analyses and graphing were conducted using STATA version 15.1 (StataCorp, College Station, USA). The dose of PQ was categorised according to the daily dose as low (<0.375 mg/kg/day), intermediate (0.375 -<0.75 mg/kg/day) or high (≥0.75 mg/kg/day). Continuous variables are presented as median, range and interquartile range (IQR). Categorical variables are presented as number and proportion. Qualitative G6PD testing was assumed to detect deficiency in patients with less than 30% enzyme activity. The risks of SAEs reported in clinical trials were stratified *a priori* into three groups: patients with unknown G6PD status, patients with G6PD activity >30%, and females with G6PD activity >30%.

### Ethics

Original studies received ethical approval via the relevant approval authorities. Ethical approval for this systematic review was obtained from the Human Research Ethics Committee of the Northern Territory Department of Health and Menzies School of Health Research (HREC reference number: 2020-3935).

#### Box 1

Classification of certainty of the diagnosis of PQ-associated severe haemolysis

**<u>Probable</u>**

**Paired Hb recorded:**

1. Fall in Hb to < 7g/dL and >25% fractional fall
2. Fall in Hb ≥5g/dL
3. (Fall in Hb 3-4.99 g/dL or >25% fall) AND (dark, brown or red urine or jaundice or shortness of breath or heart failure or blood transfusion or renal replacement therapy or death)

**Single Hb recorded after PQ:**

- Hb <7g/dL AND (dark, brown or red urine or jaundice) AND (shortness of breath or heart failure or blood transfusion or renal replacement therapy or death)

**<u>Possible</u>**

**No Hb recorded:**

- (dark urine or jaundice) AND (blood transfusion or renal replacement therapy or death)

**Single Hb recorded after PQ:**

- Hb <7g/dL AND (dark, brown, or red urine or jaundice or shortness of breath or heart failure or blood transfusion or renal replacement therapy or death)
- Hb ≥7 g/dL AND (dark urine or jaundice) AND (shortness of breath or heart failure or blood transfusion or renal replacement therapy or death)

**<u>Uncertain</u>**

- Not fulfilling probable or possible criteria but considered serious PQ-associated haemolysis by authors

Hb = haemoglobin; PQ - primaquine

Paired Hb = Hb reported before and after PQ administration Single Hb = Hb reported only after PQ administration

All cases required temporal association with PQ within the preceding 4 weeks.

## Results

### First review - serious adverse events reported in clinical trials

Between January 1990 and August 2021, 249 published *P. vivax* clinical trials (428 treatment arms) were identified, of which 138 (55.4%) fulfilled the inclusion criteria and were reviewed for SAEs (Figure 1). 70 trials (124 treatment arms, 16,214 patients; 44 (62.9%) randomised studies) explicitly mentioned adverse events and collectively included 9,824 patients treated with PQ. A further 6,390 patients were not treated with PQ (table 1).

**Figure 1.**
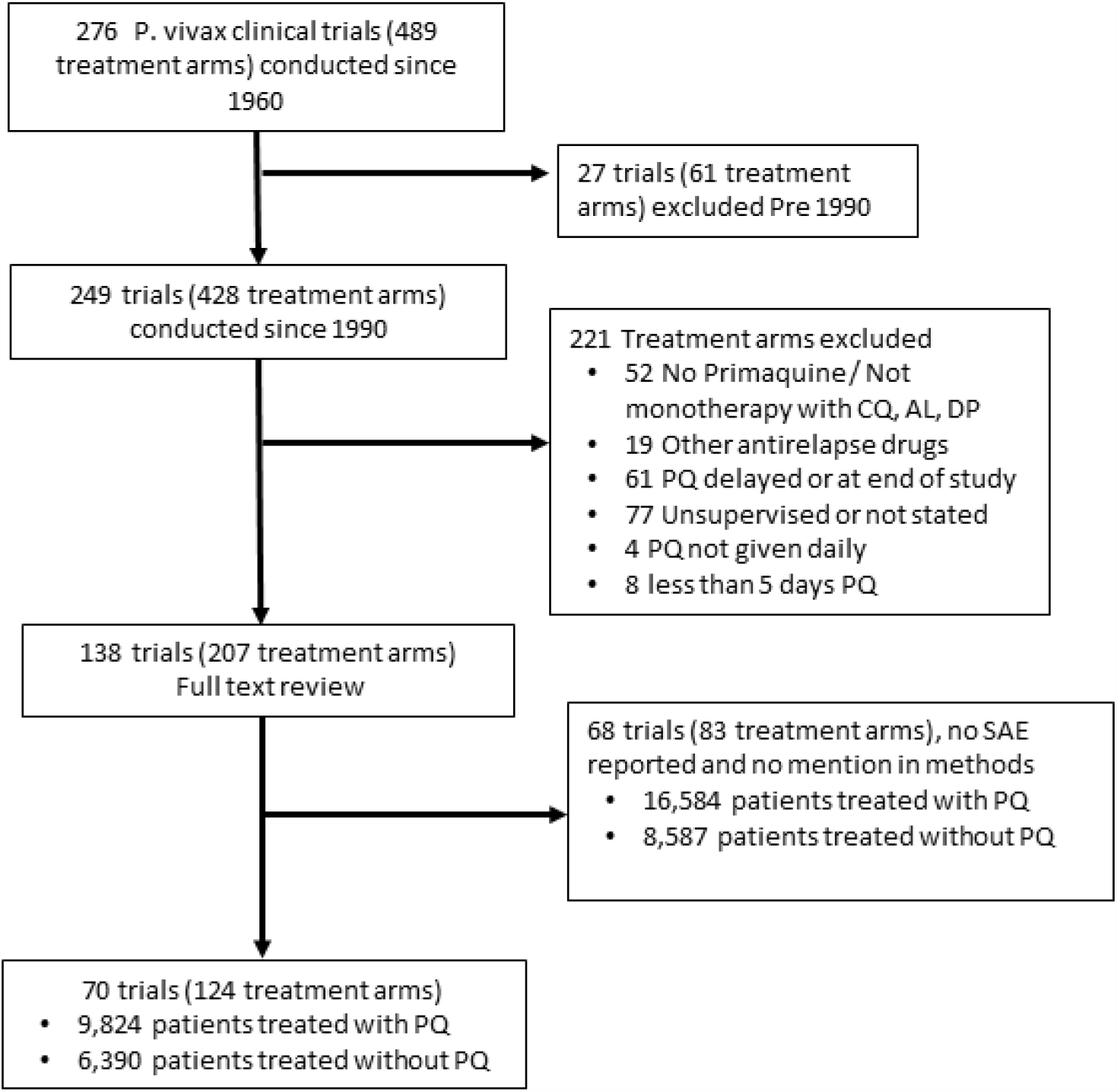
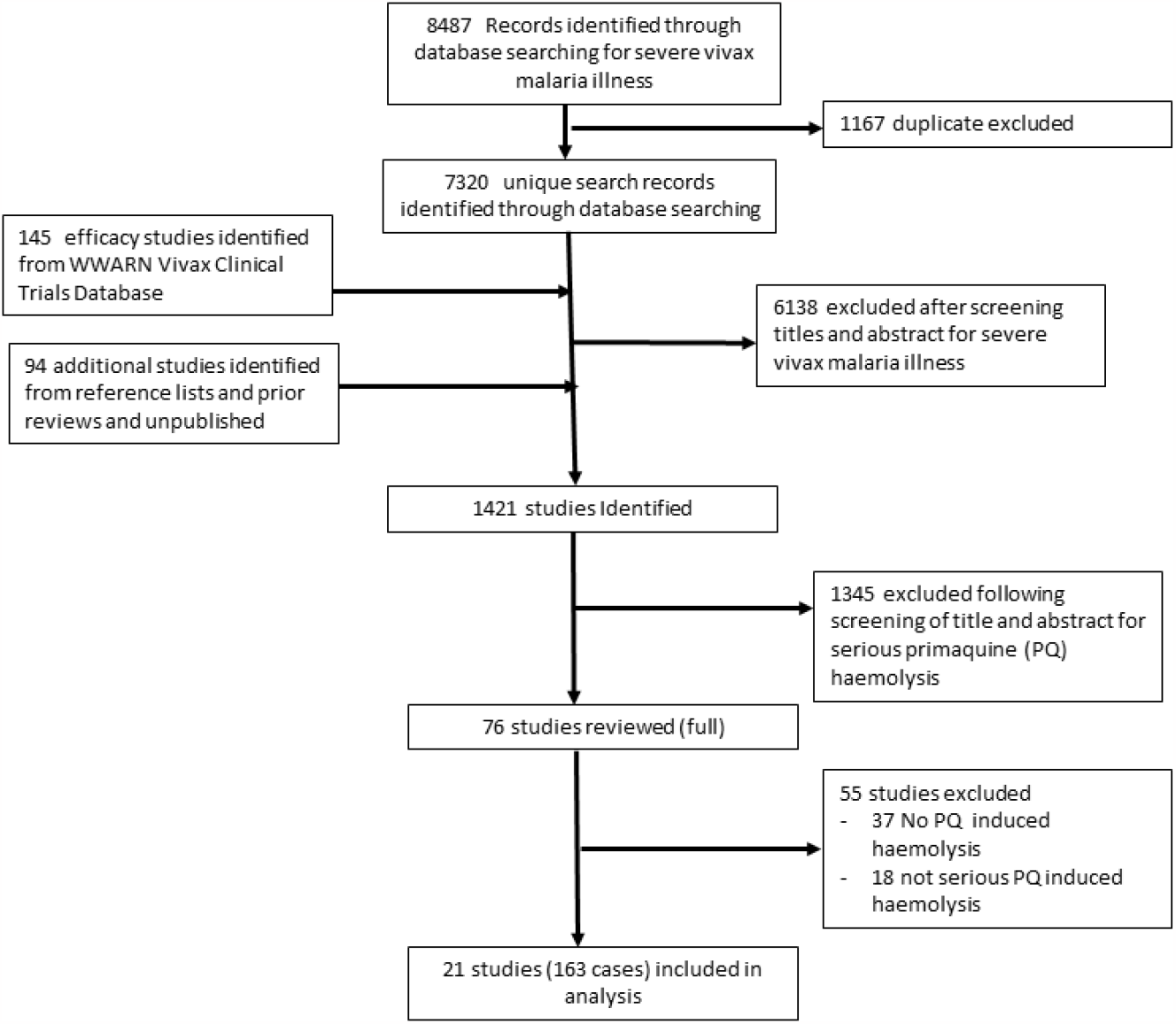
Data extraction process for A) Review of SAEs in clinical trials and B) Review of severe PQ-associated haemolysis.

**Table 1.**
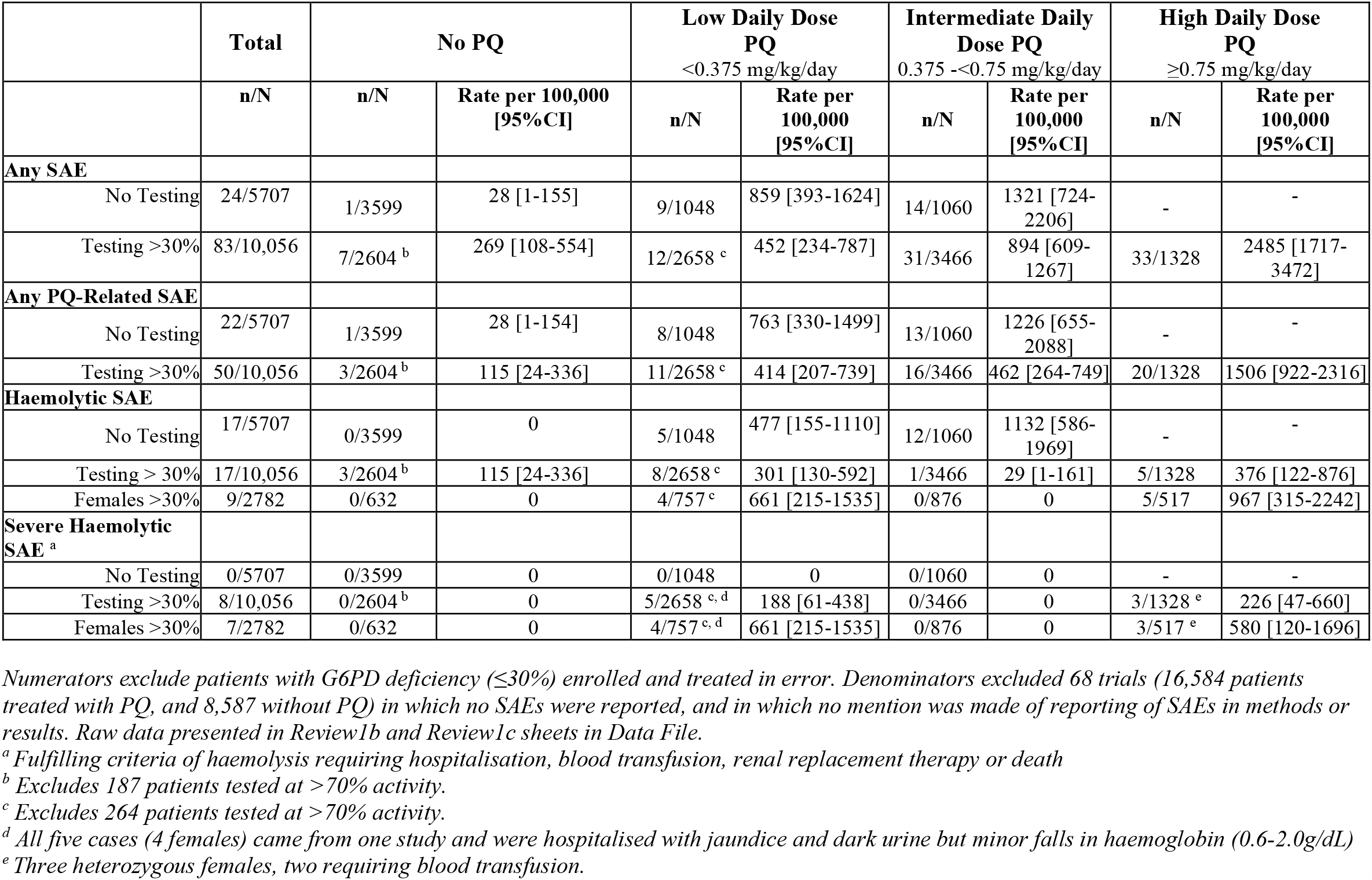
Risks of serious adverse events reported in clinical trials.

|  | Total | No PQ |  | Low Daily Dose<br>PQ<br><0.375 mg/kg/day |  | Intermediate Daily<br>Dose PQ<br>0.375 -<0.75 mg/kg/day |  | High Daily Dose<br>PQ<br>≥0.75 mg/kg/day |  |
| --- | --- | --- | --- | --- | --- | --- | --- | --- | --- |
|  | n/N | n/N | Rate per 100,000<br>[95%CI] | n/N | Rate per<br>100,000<br>[95%CI] | n/N | Rate per<br>100,000<br>[95%CI] | n/N | Rate per<br>100,000<br>[95%CI] |
| **Any SAE** |  |  |  |  |  |  |  |  |  |
| No Testing | 24/5707 | 1/3599 | 28 [1-155] | 9/1048 | 859 [393-1624] | 14/1060 | 1321 [724-2206] | - | - |
| Testing >30% | 83/10,056 | 7/2604<sup>b</sup> | 269 [108-554] | 12/2658<sup>c</sup> | 452 [234-787] | 31/3466 | 894 [609-1267] | 33/1328 | 2485 [1717-3472] |
| **Any PQ-Related SAE** |  |  |  |  |  |  |  |  |  |
| No Testing | 22/5707 | 1/3599 | 28 [1-154] | 8/1048 | 763 [330-1499] | 13/1060 | 1226 [655-2088] | - | - |
| Testing >30% | 50/10,056 | 3/2604<sup>b</sup> | 115 [24-336] | 11/2658<sup>c</sup> | 414 [207-739] | 16/3466 | 462 [264-749] | 20/1328 | 1506 [922-2316] |
| **Haemolytic SAE** |  |  |  |  |  |  |  |  |  |
| No Testing | 17/5707 | 0/3599 | 0 | 5/1048 | 477 [155-1110] | 12/1060 | 1132 [586-1969] | - | - |
| Testing > 30% | 17/10,056 | 3/2604<sup>b</sup> | 115 [24-336] | 8/2658<sup>c</sup> | 301 [130-592] | 1/3466 | 29 [1-161] | 5/1328 | 376 [122-876] |
| Females >30% | 9/2782 | 0/632 | 0 | 4/757<sup>c</sup> | 661 [215-1535] | 0/876 | 0 | 5/517 | 967 [315-2242] |
| **Severe Haemolytic SAE<sup>a</sup>** |  |  |  |  |  |  |  |  |  |
| No Testing | 0/5707 | 0/3599 | 0 | 0/1048 | 0 | 0/1060 | 0 | - | - |
| Testing >30% | 8/10,056 | 0/2604<sup>b</sup> | 0 | 5/2658<sup>c, d</sup> | 188 [61-438] | 0/3466 | 0 | 3/1328<sup>e</sup> | 226 [47-660] |
| Females >30% | 7/2782 | 0/632 | 0 | 4/757<sup>c, d</sup> | 661 [215-1535] | 0/876 | 0 | 3/517<sup>e</sup> | 580 [120-1696] |

*Numerators exclude patients with G6PD deficiency (≤30%) enrolled and treated in error. Denominators excluded 68 trials (16,584 patients* *treated with PQ, and 8,587 without PQ) in which no SAEs were reported, and in which no mention was made of reporting of SAEs in methods or* *results. Raw data presented in Review1b and Review1c sheets in Data File.*
<sup>a</sup> *Fulfilling criteria of haemolysis requiring hospitalisation, blood transfusion, renal replacement therapy or death*
<sup>b</sup> *Excludes 187 patients tested at >70% activity.*
<sup>c</sup> *Excludes 264 patients tested at >70% activity.*
<sup>d</sup> *All five cases (4 females) came from one study and were hospitalised with jaundice and dark urine but minor falls in haemoglobin (0.6-2.0g/dL)*
<sup>e</sup> *Three heterozygous females, two requiring blood transfusion.*

A total of 110 SAEs were reported, of which 68 (67.3%) were attributed by the authors to PQ treatment. There were 37 episodes of acute haemolysis; 34 occurred in patients treated with PQ and 3 in patients treated without PQ. Three haemolytic SAEs occurred in patients with G6PD deficiency (<30%) who had been treated with PQ in error.

The reasons stated for classification as a haemolytic SAE included 16 patients who had a >2.5 g/dL drop in Hb or >25% fall (in all cases PQ was continued without interruption), 5 patients who had a >3 g/dL drop in Hb or >30% fall (PQ was continued in 4 cases), 3 patients who had dark urine, and 6 patients who had a fall in Hb to below 7g/dL (3 required blood transfusion and in two of the remaining cases PQ was temporarily ceased and restarted). A further 6 patients, all in the same study, were hospitalised for management, although their Hb nadir ranged from 8.5 to 13.2 g/dL, with a maximum absolute fall in Hb of 2 g/dL. One patient was reported as having acute haemolysis with jaundice (total bilirubin 105 µmol/L) but no further details could be ascertained. Overall, 27.2% (10/37) of the reported haemolytic SAEs were classified by us as ‘severe’ based on the need for hospitalisation, blood transfusion, renal replacement therapy or death.

The risks of SAE and haemolysis in patients with unknown G6PD status or G6PD activity ≥30% are presented in Table 1. The overall incidence of PQ-associated haemolytic SAE (all PQ dosing regimens) was 188 [95% confidence interval (95%CI) 103-315] per 100,000 and the overall risk of severe PQ haemolysis was 107 [95%CI 46-212] per 100,000.

### Second Review - Reports of Severe Haemolysis

In the second review, a total of 8,487 articles were identified describing patients with vivax malaria requiring admission to hospital. After removal of 1,167 duplicate publications, addition of a further 239 articles (identified from clinical trials and previous reviews) and exclusion of articles after screening of the titles and abstracts, a total of 76 articles remained for full-text review (figure 1). 55 studies were subsequently excluded because they did not report patients fulfilling the criteria for severe PQ-associated haemolysis. A total of 163 individuals with severe PQ-associated haemolysis were identified from 21 articles published between 1953 and 2020: 43 were from Asia (13 articles), 116 from South America (5 articles), 3 from North America (2 articles) and 1 from Oceania (1 article) (Figure 1). 17 patients (8 articles) were reported in randomised controlled trials and 146 patients (13 articles) were reported in case reports. A summary of the studies included in the analysis is provided in the Supplementary Data File.

The median age of individuals with severe PQ-associated haemolysis was 18 (IQR: 12 - 28; range 1 – 84) years and 132 (80.9%) were males. Overall, 134 (82.2%) patients had *P. vivax* infection and 6 (3.7%) had mixed *P. vivax* and *P. falciparum* infection. A further 23 individuals (14.1%) were negative when tested for malaria, 19 of whom were presumed by the authors to have had previous *P. vivax* infection. Of the 140 individuals treated for acute malaria, 135 (96.4%) also received chloroquine, 4 (2.9%) dihydroartemisinin-piperaquine and 1 (0.7%) amodiaquine.

The duration of the PQ regimen prescribed was reported in 156 cases. Eighteen (11.5%) were prescribed 14 days of treatment, one (0.6%) 8 days treatment, 121 (77.6%) 7 days treatment and 16 (10.3%) 5 days or less. Two (1.3%) patients were prescribed weekly PQ for eight weeks. The target daily PQ doses prescribed are provided in Table 2. The total dose prescribed was <2.5 mg/kg for 16 (10.3%) patients, 2.5-5 mg/kg for 132 (84.6%) patients and ≥5 mg/kg for 8 (5.1%) patients.

**Table 2.** Characteristics and clinical outcomes of individuals with severe PQ-associated haemolysis.

| Clinical manifestations of severe haemolysis | Probable PQ-associated<br>N=48 |  | Possible PQ-associated<br>N=53 |  | Uncertain PQ-associated<br>N=62 |  |
| --- | --- | --- | --- | --- | --- | --- |
|  | N | n (%) | N | n (%) | N | n (%) |
| **Symptoms & signs** |  |  |  |  |  |  |
| Dark urine | 36 | 29 (80.6) | 38 | 34 (89.5) | 39 | 38 (97.4) |
| Pallor | 40 | 38 (95.0) | 35 | 31 (88.6) | 40 | 36 (90.0) |
| Dyspnoea | 33 | 9 (27.3) | 26 | 7 (26.9) | 30 | 2 (6.7) |
| Dizziness | 17 | 9 (52.9) | 9 | 4 (44.4) | 8 | 3 (37.5) |
| Fever | 38 | 24 (63.2) | 31 | 16 (51.6) | 32 | 7 (21.9) |
| Severe fatigue | 20 | 13 (65.0) | 6 | 5 (83.3) | 3 | 2 (66.7) |
| Heart failure | 31 | 3 (9.7) | 31 | 6 (19.4) | 33 | 0 (0.0) |
| Chest pain | 7 | 1 (14.3) | 3 | 0 (0.0) | 0 | 0 (0.0) |
| Jaundice | 44 | 39 (88.6) | 48 | 44 (91.7) | 54 | 47 (87) |
| Low urine output | 14 | 2 (14.3) | 4 | 0 (0.0) | 5 | 2 (40) |
| **Laboratory findings** |  |  |  |  |  |  |
| Glucose 6-Phosphate dehydrogenase |  |  |  |  |  |  |
| Normal | 47 | 0 (0.0) | 51 | 2 (3.9) | 61 | 5 (8.2) |
| Intermediate | 47 | 2 (4.4) | 51 | 0 (0.0) | 61 | 0 (0.0) |
| Deficient | 47 | 45 (95.7) | 51 | 49 (96.1) | 61 | 56 (91.8) |
| Total bilirubin, >1 mg/dL | 37 | 31 (83.8) | 38 | 35 (92.1) | 52 | 46 (88.5) |
| Indirect bilirubin, >0.8 mg/dL | 36 | 30 (83.3) | 36 | 33 (91.7) | 43 | 37 (86.1) |
| Urine dipstick blood, positive | 21 | 10 (47.6) | 27 | 19 (60.4) | 31 | 21 (67.7) |
| Urine dipstick urobilinogen, positive | 20 | 3 (15) | 25 | 14 (66) | 30 | 12 (40) |
| White blood cell count, >12,000/μL | 32 | 20 (62.5) | 38 | 20 (52.6) | 36 | 10 (27.8) |
| **Target daily dose of PQ** |  |  |  |  |  |  |
| 0.25 mg/kg/day | 47 | 5 (10.6) | 52 | 12 (23.1) | 57 | 14 (24.6) |
| 0.5 mg/kg/day | 47 | 36 (76.6) | 52 | 39 (75.0) | 57 | 43 (75.4) |
| 0.75 mg/kg/day | 47 | 2 (4.3) | 52 | 0 (0.0) | 57 | 0 (0.0) |
| 1.0 mg/kg/day | 47 | 4 (8.5) | 52 | 1 (1.9) | 57 | 0 (0.0) |
| **Management** |  |  |  |  |  |  |
| Blood transfusion | 48 | 45 (93.8) | 42 | 32 (76.2) | 43 | 0 (0.0) |
| Renal replacement therapy | 39 | 4 (10.3) | 41 | 4 (9.8) | 42 | 0 (0.0) |
| Hospitalised (no blood transfusion or renal replacement therapy) | 3 | 3 (100) | 19 | 19 (100) | 62 | 62 (100) |
| **Outcome** |  |  |  |  |  |  |
| Died | 48 | 0 (0.0) | 53 | 7 (13.2) | 62 | 0 (0.0) |

*N = number of patients for whom data were available. n= number of patients with manifestation present*

**Table 3.** Clinical manifestations and outcomes of patients with probable or possible severe PQ-associated haemolysis.

|  | **Blood transfusion**<br>**N=76** |  | **Renal replacement therapy**<br>**N=8** |  | **Death**<br>**N=7** |  | **All patients**<br>**N=101** |  |
| --- | --- | --- | --- | --- | --- | --- | --- | --- |
|  | **N** | **n (%)** | **N** | **n (%)** | **N** | **n (%)** | **N** | **n (%)** |
| **Clinical manifestations** |  |  |  |  |  |  |  |  |
| Dark urine | 57 | 48 (84.2) | 5 | 5 (100) | 4 | 4 (100) | 74 | 63 (85.1) |
| Pallor | 60 | 56 (93.3) | 8 | 7 (87.5) | 2 | 2 (100) | 75 | 69 (92.0) |
| Dyspnoea | 47 | 8 (17.0) | 7 | 2 (28.6) | 3 | 3 (100) | 59 | 16 (27.1) |
| Dizziness | 22 | 11 (50.0) | 4 | 1 (25.0) | 0 | 0 (0.0) | 26 | 13 (50.0) |
| Fever | 58 | 37 (63.8) | 7 | 4 (57.1) | 1 | 1 (100) | 69 | 40 (57.8) |
| Severe fatigue | 21 | 15 (71.4) | 0 | 0 (0.0) | 0 | 0 (0.0) | 26 | 18 (69.2) |
| Heart failure | 51 | 7 (13.7) | 6 | 6 (100) | 0 | 0 (0.0) | 62 | 9 (14.5) |
| Chest pain | 8 | 1 (12.5) | 8 | 0 (0.0) | 0 | 0 (0.0) | 10 | 1 (10.0) |
| Jaundice | 67 | 59 (88.1) | 7 | 7 (100) | 6 | 6 (100) | 92 | 83 (90.2) |
| Low urine output | 14 | 2 (14.3) | 3 | 2 (66.7) | 0 | 0 (0.0) | 18 | 2(11.1) |
| **Laboratory findings** |  |  |  |  |  |  |  |  |
| Haemoglobin >3g/dl drop | 11 | 9 (81.8) | 0 | 0 (0.0) | 0 | 0 (0.0) | 14 | 12 (85.7) |
| Haemoglobin >25% fall | 11 | 11 (100) | 0 | 0 (0.0) | 0 | 0 (0.0) | 14 | 14 (100) |
| Total bilirubin > 1 mg/dL | 59 | 51 (86.4) | 7 | 5 (71.4) | 1 | 1 (100) | 75 | 66 (88.0) |
| Indirect bilirubin > 0.8 mg/dL | 59 | 51 (86.4) | 7 | 5 (71.4) | 1 | 1 (100) | 72 | 63 (87.5) |
| Urine dipstick blood, positive | 31 | 17 (54.8) | 6 | 3 (50.0) | 5 | 5 (100) | 48 | 29 (60.4) |
| Urobilinogen dipstick positive | 30 | 5 (16.7) | 5 | 1 (20.0) | 3 | 3 (100) | 45 | 17 (37.8) |
| White blood cell count > 12,000/μL | 56 | 33 (58.9) | 7 | 4 (57.1) | 3 | 3 (100) | 70 | 40 (57.1) |

*N = number of patients in whom data was available. n= number of patients with manifestation present*
*Patients could present with more than one outcome thus total number of patients does not equal summation of those who received blood transfusion, renal replacement therapy, or who died.*

G6PD status was recorded for 159 (97.5%) of the 163 patients, with a qualitative assay used in 145 (91.2%) patients and a quantitative assay in 10 (6.3%) patients. The assay method was not stated in 4 (2.5%) cases. Overall, 150 (94.3%) individuals were categorised as G6PD deficient and two (1.3%) had intermediate G6PD deficiency. Seven (4.4%) patients were G6PD normal according to a qualitative G6PD test (assumed to have ≥30% activity), four of whom were female. The certainty of the diagnosis of severe PQ-associated haemolysis was categorised as probable in 48 (29.5%) cases, possible in 53 (32.5%) cases and uncertain in 62 (38.0%) cases. G6PD activity was documented in 94 of the patients with probable or possible severe haemolysis, of whom 96.8% (n=91) were G6PD deficient (<30% activity) and 3 were heterozygous females (64%, 94% and >30% enzyme activity respectively).

### Clinical presentation and management of patients

All subsequent results are restricted to the 101 patients with probable or possible severe PQ-related haemolysis, 95 of whom were G6PD deficient (<30%), (three patients were heterozygous females and three patients did not have a reported G6PD status). The most common clinical presentations of haemolysis were pallor (92.0%, 69/75), dark urine (85.1%, 63/74) and jaundice (90.2%, 83/92) (table 2). Fourteen patients had a haemoglobin measurement before and after initiation of PQ. In these patients the median fall in haemoglobin was 4.4 g/dL (IQR: 3.7 - 6.6; Range: 2.2 - 8.9). Of the patients in whom serum bilirubin was measured, 88% (66/75) had raised total bilirubin concentration (median = 4 mg/dL, interquartile range [IQR]: 2.1 - 5.5; range: 0.3 - 46.4). At the time that patients were hospitalised, 79.5% (58/73) of those with acute malaria were aparasitaemic, however 55.3% (26/47) were febrile and 62.5% (30/48) had a raised total white cell count.

### Timing of severe PQ-associated haemolysis

Information on the timing of manifestations of severe PQ-associated haemolysis was available for 93 (92.1%) patients, with all patients presenting within the first 7 days of commencing PQ treatment. Overall, 89.2% (83/93) developed symptoms of haemolysis within 4 days of commencing PQ. The first symptoms of haemolysis were mostly reported on day 2 or 3 (43.3%) after commencing PQ, whereas onset of severe haemolysis occurred most frequently on day 3 or 4 (55.4%) (figure 2). At the onset of manifestations of severe haemolysis, PQ had been taken for a median of 4 days (range 1-8) with a median total dose received of 2.0 mg/kg (IQR: 1.5-2.5, Range: 0.3-5.0) (figure S1 appendix pp7). In total, 82.8% (82/99) of the cases of probable or possible PQ-associated severe haemolysis occurred following administration of a daily dose ≥0.5 mg/kg (figure 3).

**Figure 2.**
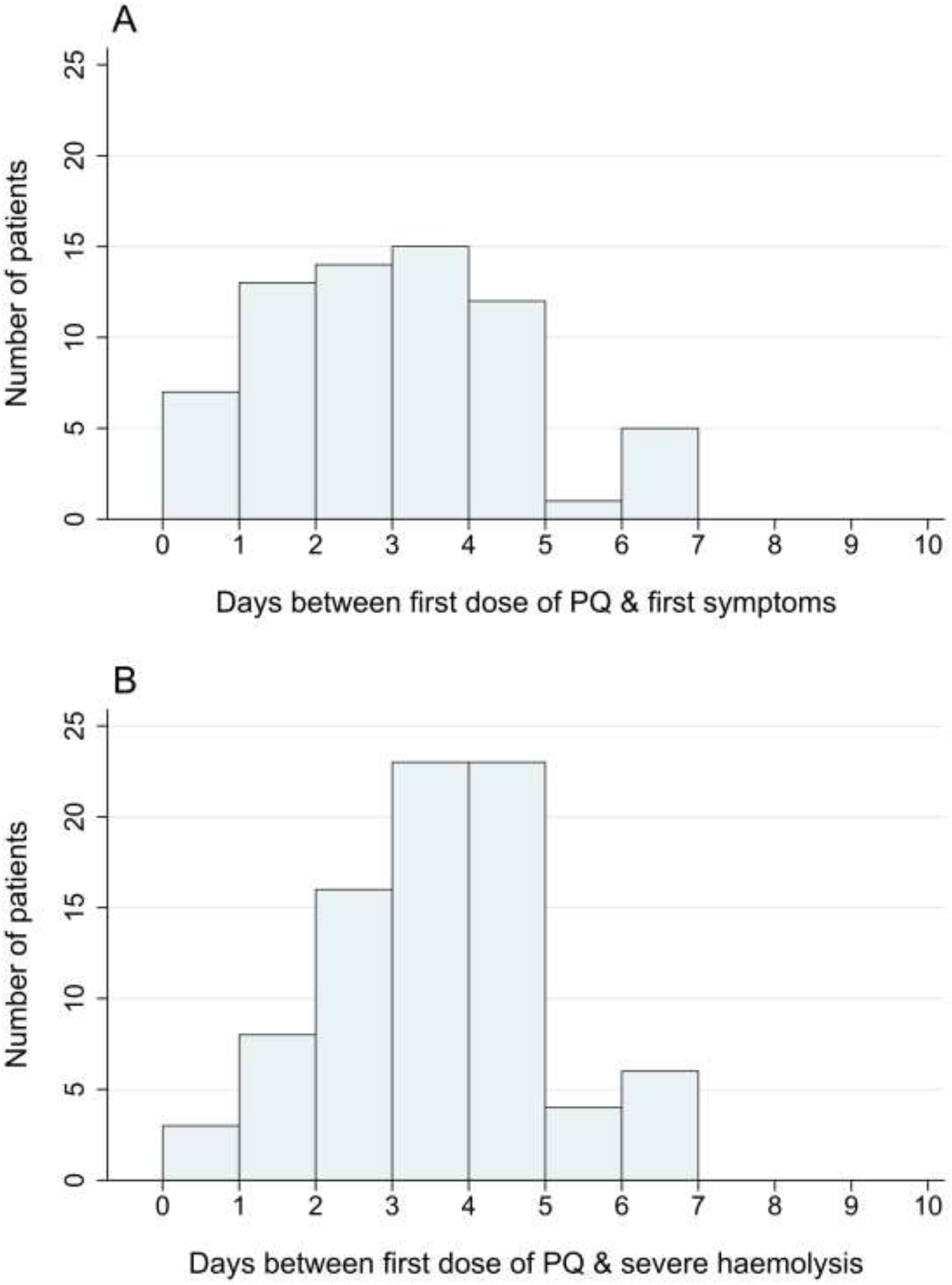
Timing of the first symptoms of haemolysis (A) and onset of severe haemolysis (B) after commencing PQ.

**Figure 3.**
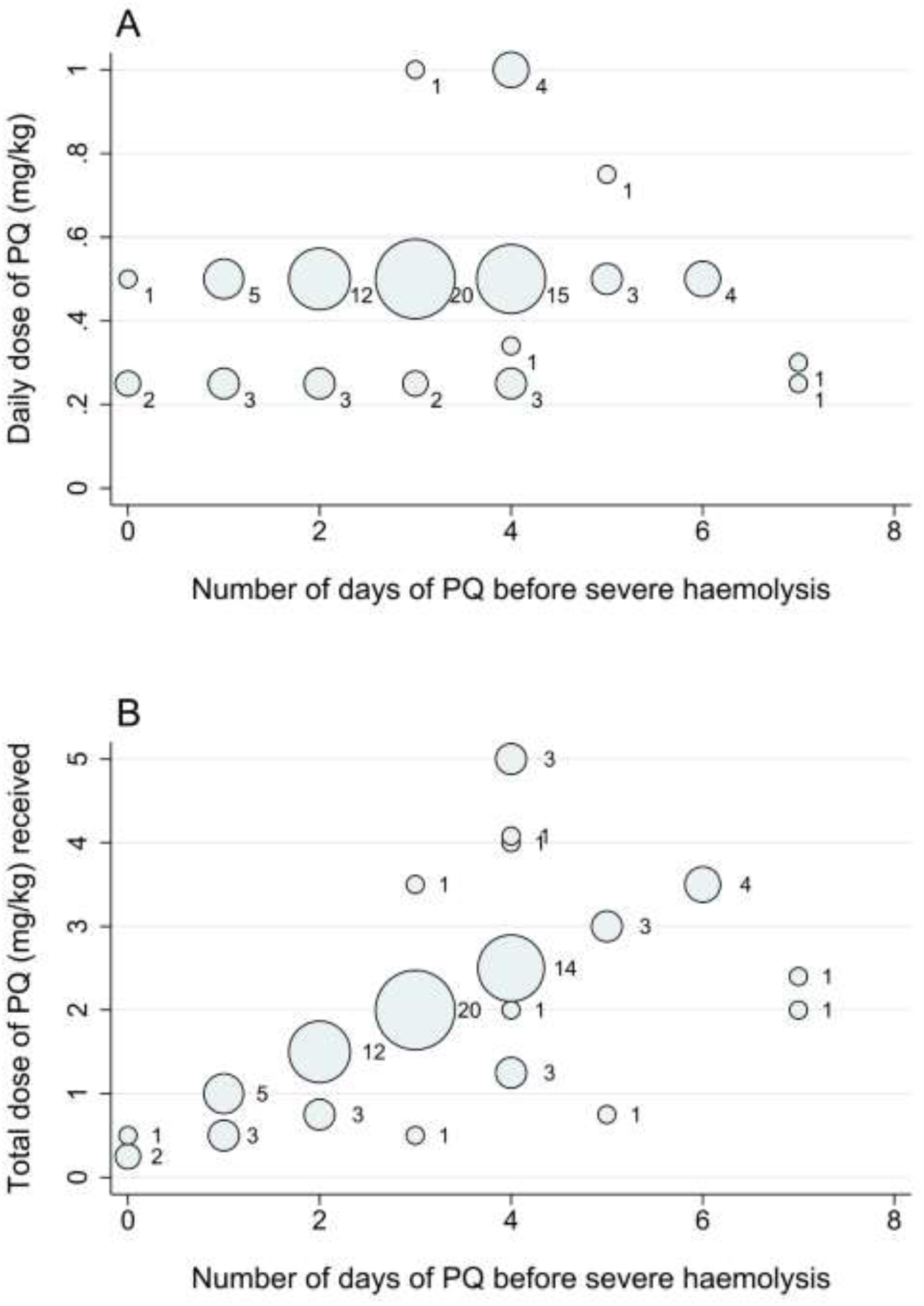
Timing of hospital admission with severe PQ-associated haemolysis according to the daily dose of PQ (A) and total PQ dose administered at that time (B). Footnote: Size and label of each dot indicates the number of events. Data restricted to 101 probable and possible cases of severe PQ-associated haemolysis

**Figure 4.**
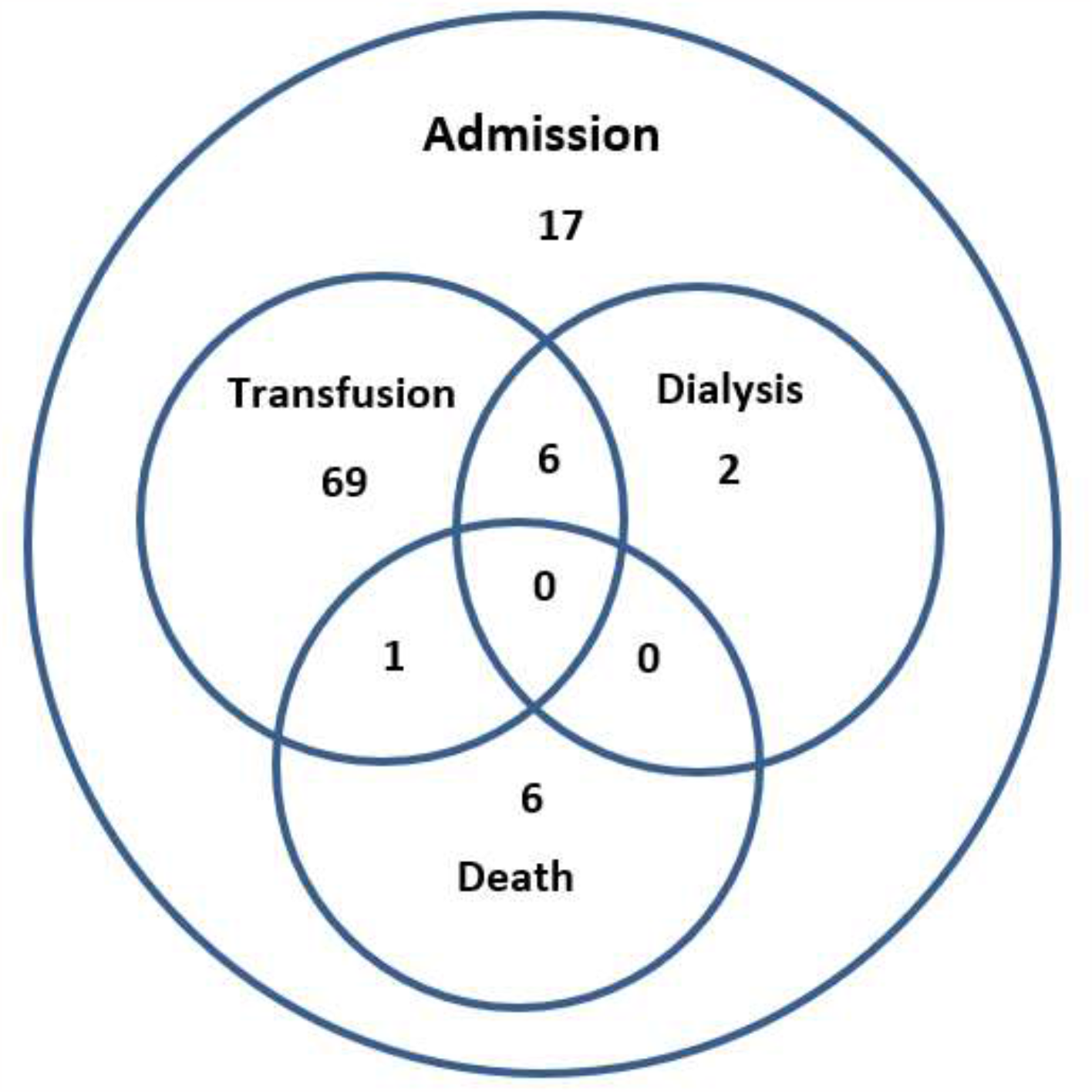
Outcomes following severe PQ-associated haemolysis. Data restricted to 101 probable and possible cases of severe PQ-associated haemolysis

### Outcomes

83.5% (76/90) of patients with severe PQ-associated haemolysis required blood transfusion and 10.3% (8/78) required renal replacement therapy. The absolute fall in Hb could not be calculated for any of the patients requiring renal replacement therapy. Seven (6.9%) patients died. All but one of the deaths occurred in males. Four deaths were reported in Sri Lankan children, all of whom received PQ for radical cure of *P. vivax*. The cause of death was reported as cardiac failure from anaemia in three of these patients and irreversible renal failure in one. Three deaths were reported in Brazilian patients, all males. Overall, 5 of the 7 patients who died were reported to be G6PD deficient, with the G6PD status unknown in the remaining two cases. One of the patients who died received a blood transfusion, but information on blood transfusion or renal replacement therapy was not documented for the remaining 6 patients. Of the patients who survived, 72.7% (56/77) were discharged from hospital within one week of admission.

## Discussion

Radical cure of *P. vivax* malaria requires treatment of both blood and liver stages of the parasite to ensure clinical recovery and reduce the risk of subsequent relapse. Without radical cure, relapses occur in up to 70% of patients, the risk and frequency of which vary with host immunity, background transmission intensity and geographical location.^15^ The antirelapse efficacy of PQ is related to the total dose of PQ administered, whereas for the 7- and 14-day regimens the risk of haemolysis is related to the daily dose.^16,17^ Since routine G6PD testing is often unavailable in malaria endemic countries, most national malaria control programs currently recommend a total dose of 3.5 mg/kg administered over 14 days, allowing the daily dose to be reduced to 0.25 mg/kg to minimise haemolysis.

Adherence to standard 14-day PQ regimens is typically poor.^18^ Seven-day regimens of higher daily doses of PQ (0.5 mg/kg) are used routinely in several endemic countries including Brazil, Peru and China,^10^ providing a total dose of 3.5 mg/kg. Recent evidence suggests that 7 mg/kg may be required to reduce the risk of recurrent parasitaemia to less than 10% in areas of both high and low relapse periodicity.^19,20^ Giving 7mg/kg over 7-as opposed to 14-days (1 mg/kg/day) is equally efficacious but the increased daily dose is associated with greater risk of SAE.^21,22^

A total of 37 haemolytic SAEs were reported in the clinical trials in our first review, 34 of which occurred following PQ administration. The criteria used to classify these adverse events as serious varied. 21 reported SAEs were based solely on a significant fall in haemoglobin (>2-3 g/dL) with all but one of these patients continuing their course of PQ to completion. In total 10 of the patients with reported SAEs required hospitalisation and blood transfusion for medical management. The overall risk of severe PQ haemolysis was estimated to be 107 per 100,000 patients with normal G6PD activity (≥30% activity) treated with PQ. The prospective clinical trials in our reviews generally applied more conservative criteria for classifying adverse events as serious (presumably with the aim of ensuring patient safety in a controlled environment) than the case reports and pharmacovigilance studies in our second review. The latter often had limited clinical and laboratory information available prior to the onset of the events. Even when comprehensive data are available, the clinical and laboratory features that justify haemolysis being classified as serious or life-threatening have yet to be defined. This results in a lack of consistent data for decision-makers to balance the risks of PQ radical cure with the risks of multiple relapses of vivax malaria in different endemic locations.^1^ Quantifying these risks will require robust definitions of haemolysis that can accommodate differential access to laboratory investigations. Standardised templates for reporting relevant clinical and laboratory details of potential haemolytic events will facilitate this process greatly. An example of a form for reporting the key data identified in our analysis is provided (appendix pp 8-12).

The second of our systematic reviews included all articles published since the introduction of PQ. The largest case series were reported from South America, where the prevalence of G6PD deficiency is high and comparatively large daily doses (0.5 mg/kg) of PQ are recommended.^8,23^ Conversely none of the cases in this review were reported from Africa, where G6PD deficiency is mostly attributable to the mild A-variant.^5^ Based on *a priori* criteria for certainty of association, 101 cases of severe haemolysis were categorised as probably or possibly related to PQ, all but one of which occurred in patients with either intermediate or severe G6PD deficiency. The first signs and symptoms of haemolysis occurred within four days of starting treatment in over 90% of cases, with hospitalisation occurring most frequently on day 4 after a median total dose of 2 mg/kg. All patients presented to hospital within the first week after starting PQ. Most cases occurred in patients receiving ≥0.5 mg/kg/day. The signs and symptoms of severe PQ-associated haemolysis in our review concurred broadly with those found in G6PD deficient patients treated with the sulphur-based antimalarial drug dapsone.^24^

A total of 12 deaths probably or possibly related to PQ-associated haemolysis have been reported in the scientific literature, of which seven followed PQ for radical cure of *P. vivax* and were included in our second review.^4^ Although other deaths have undoubtedly occurred and gone unreported, this outcome is rare, with estimates varying from 1 in 625,000 to 1 in 110,000 patients treated with PQ.^4,8^ Of the seven patients in our analysis who died, five had recorded G6PD status and were deficient at the 30% threshold.^8,25,26^ Of the 101 patients with probable or possible severe haemolysis, 98 had G6PD status recorded and all but three of these patients had G6PD deficiency with activity less than 30%. The remaining three patients were heterozygous females treated with high dose PQ (1mg/kg/day).^21^ Although there is an increased risk of haemolysis in heterozygous females with intermediate deficiency (30-70%),^27^ our findings suggest that for low (0.25 mg/kg) and intermediate (0.5mg/kg) daily doses, severe haemolysis can be minimised by a reliable qualitative test used to exclude exposure of individuals with less than 30% enzyme activity.

Widespread point-of-care testing for G6PD deficiency prior to PQ administration will facilitate exclusion of patients at greatest risk of haemolysis and reduce adverse drug reactions. However, even in optimal conditions, G6PD testing is not 100% reliable and due to logistical, financial and supply constraints, quality tests may not always be available. In these cases the risks of PQ-associated haemolysis need to be balanced against the cumulative risks of recurrent episodes of parasitaemia and associated morbidity and mortality.^1,28^ In this context it is critical to understand the clinical presentation and outcomes of severe PQ-associated haemolysis, so that patients with impending haemolysis can be identified and mitigating strategies taken to avoid further deterioration.

PQ-associated haemolysis is most prominent during the first week of treatment, as circulating senescent erythrocytes with low G6PD activity and greatest vulnerability to oxidant stress are cleared.^27^ With time, increased production of relatively G6PD replete reticulocytes partially compensates for the increased red cell loss and increases the median tolerance of circulating erythrocytes to oxidative stress.^29^ This presumably explains the lack of new presentations with severe haemolysis beyond one week of starting PQ treatment. Hospitalisation was delayed with respect to symptom onset by one to two days suggesting that there is an opportunity to prevent severe manifestations of haemolysis with early intervention. Based on these observations, routine clinical review within 5 days of starting PQ would identify most patients at risk of severe haemolysis and allow early cessation of PQ to prevent further clinical deterioration.

Strengths of this paper include the use of consistent and strict *a priori* definitions for severe haemolysis and categorisation of likelihood of a causal association with PQ. Individualised patient data were used and authors approached for additional unpublished information. Despite this, clinical information available for many patients was insufficient to enable a full assessment of likely causality. Our assessment of the frequency of severe PQ-associated haemolysis will have been subject to ascertainment bias as many cases will not have been formally reported in the medical literature. Conversely, despite our attempts to define a causal association with drug administration, some haemolytic events may have been inadvertently attributed to PQ. A previous pooled analysis has shown that approximately 1 in 1,000 patients with *P. vivax* malaria treated without PQ had acute falls in haemoglobin >5 g/dL,^15^ remarkably similar to the risk of severe haemolysis we estimated in patients with >30% enzyme activity (107 per 100,000). In our series, four G6PD deficient patients developed severe haemolysis on the day of initial presentation within hours of receiving the first dose of PQ. It is likely that these cases were attributable to acute malaria rather than drug administration.

In conclusion, severe PQ-associated haemolysis with clinical decompensation requiring urgent medical intervention is rare but when it does occur, the first signs tend to become apparent within 2 to 3 days of starting treatment. Routine clinical review within 5 days to screen for symptoms such as dark urine, jaundice, pallor, severe fatigue and fever would facilitate early detection of the majority of cases of impending severe haemolysis enabling remedial action to be taken to prevent adverse outcomes. Diverse criteria have been used to categorise haematological SAEs and this limits comparisons of their frequency between different locations and dosing regimens. Standardised definitions would facilitate valid pharmacovigilance in clinical trials and real-world settings. Whilst widely available point-of-care testing for G6PD deficiency will be critical for reducing the risk of severe adverse drug reactions in vulnerable patients, severe haemolysis may still occur due to parasite-induced haemolysis, idiosyncratic reactions and diagnostic errors. Hence any public health intervention must include community engagement to ensure patients and healthcare providers are aware of the haemolytic risks of malaria and its treatment and the need to represent for medical review in case of clinical deterioration.

## Data Availability

Data are contained in the supplementary data file.

## Acknowledgements

We thank the investigators of several reports for providing additional information on serious adverse events, including André Daher and Kevin Baird.

## Contributors

DY, EG, RC, RP and ND conceptualised the study. All authors were responsible for data collection and curation. DY, RP, RC and ND analysed the data. All authors helped interpret the data. DY, RP and ND wrote the original draft of the paper. All authors reviewed and edited the paper.

## Research in Context

### Evidence before this study

Primaquine has been used since the 1950s for preventing relapses of *Plasmodium vivax* malaria but has the potential to cause severe haemolysis, particularly in patients with glucose-6-phosphate dehydrogenase (G6PD) deficiency. We did two systematic reviews to examine the definitions, frequency, timing, clinical nature, management and outcomes of primaquine-induced haemolysis. In the first of two reviews, we examined all studies in the WorldWide Antimalarial Resistance Network (WWARN) “vivax surveyor” repository (previously registered at PROSPERO) and extracted all clinical trials published between January 1, 1990 and August 23, 2021 in which patients with *Plasmodium vivax* malaria were treated with either partial or fully supervised primaquine therapy commencing within 7 days of blood schizontocidal therapy and lasting for at least 5 days’ duration. Studies not reporting adverse events were excluded. In the second review, we searched PubMed, Web of Science, Embase and the Cochrane Central Database without language restrictions from January 1, 1940 to May 20, 2020 for studies reporting severe primaquine-associated haemolysis using the terms vivax and (hospital* or renal or dialysis or transfusion or severe or serious or haemolysis or hemolysis or fatal or death or died or methemoglobin* or methaemoglobin* or ‘cerebral complicat*’ or convuls* or unconscious* or prostrat* or ‘kidney injury’ or ‘renal failure’ or ‘renal impairment’ or hemoglobinuria or haemoglobinuria or ‘circulatory collapse’ or shock or jaundice or hyperbilirubinemia or hyperbilirubinaemia or ‘hepatic dysfunction’ or ‘liver dysfunction’ or bleeding or hemorrhage or haemorrhage or thrombocytopenia or thrombocytopaenia or ‘disseminated intravascular coagulation’ or DIC or ‘acute respiratory distress syndrome’ or ARDS or ‘pulmonary edema’ or ‘pulmonary oedema’ or ‘metabolic acidosis’ or hyperlactat* or ‘severe anemia’ or ‘severe anaemia’ or hypoglycemia or hypoglycaemia or complicat*). We also searched the WWARN repository of *P. vivax* clinical trials as well as conference proceedings and reference lists of identified articles. The first review yielded 70 trials (124 treatment arms, 9,824 patients treated with primaquine) and the second review yielded 21 studies reporting individualised data on patients with potential primaquine-associated haemolysis. National and subnational guidelines for primaquine radical cure of vivax malaria vary markedly in different locations because of differences in the perceived risk of primaquine-associated haemolysis and adverse consequences of vivax malaria relapses. Some countries and territories omit primaquine altogether. There is no standardized definition of serious or life-threatening primaquine-associated haemolysis, making comparisons of the frequency of this feared outcome across different endemic locations difficult.

### Added value of this study

Our first review provides the first population-level estimate of the frequency of severe primaquine-associated haemolysis requiring hospitalisation or blood transfusion of 107 per 100,000 patients with normal G6PD activity treated with primaquine for vivax malaria. It also highlights a wide spectrum of definitions used to classify haemolytic events as serious, many of which did not have important attendant clinical outcomes. Our second review provides an exhaustive summary of severe primaquine-associated haemolytic events reported in the medical literature. In total there have been 163 cases requiring hospitalisation for haemolysis after primaquine for vivax malaria and 7 deaths, 5 of which occurred in patients with known G6PD deficiency. Our review demonstrates that the first symptoms or signs of impending severe haemolysis typically occur on day two or three of treatment and that need for hospitalisation is delayed by a further one to two days.

### Implications of all the available evidence

Severe primaquine-associated haemolysis is very rare but can be fatal. Since need for hospitalisation is typically delayed with respect to symptom onset, clinical review within 5 days of initiation of treatment could facilitate detection of impending severe haemolysis and enable remedial action. Definitions of serious primaquine-associated haemolysis need to be standardised so that rates of haemolysis can be compared across different endemic locations. A practicable point-of-care test for G6PD deficiency would increase the safety of primaquine use but would not negate the need for improved, standardised pharmacovigilance.

