## Supplementary appendix file for "Severe haemolysis during primaquine radical cure of *Plasmodium vivax* malaria: two systematic reviews and individual patient data descriptive analyses"

### Table of Contents

*Table of Contents*...1

*PRIMSA – IPD Checklist* ...2

*Box 1 - Search strategy for Review 1 of P. vivax Antimalarial Clinical Trials*...5

*Box 2 - Search strategy for Review 2 of Severe PQ-associated Haemolysis* ...6

*Figure S1 - Total dose of PQ (mg/kg) administered (A) before first symptoms of haemolysis and (B) manifestation of severe haemolysis* ...7

*Example of Form for reporting drug induced haemolysis following treatment of malaria* ...8

### PRIMSA – Checklist

| Section/topic | # | Checklist item | Reported on page # |
|---|---|---|---|
| **TITLE** | | | |
| Title | 1 | Identify the report as a systematic review, meta-analysis, or both. | 1 |
| **ABSTRACT** | | | |
| Structured summary | 2 | Provide a structured summary including, as applicable: background; objectives; data sources; study eligibility criteria, participants, and interventions; study appraisal and synthesis methods; results; limitations; conclusions and implications of key findings; systematic review registration number. | 3-4 |
| **INTRODUCTION** | | | |
| Rationale | 3 | Describe the rationale for the review in the context of what is already known. | 5-6 |
| Objectives | 4 | Provide an explicit statement of questions being addressed with reference to participants, interventions, comparisons, outcomes, and study design (PICOS). | 6 |
| **METHODS** | | | |
| Protocol and registration | 5 | Indicate if a review protocol exists, if and where it can be accessed (e.g., Web address), and, if available, provide registration information including registration number. | 6-7 |
| Eligibility criteria | 6 | Specify study characteristics (e.g., PICOS, length of follow-up) and report characteristics (e.g., years considered, language, publication status) used as criteria for eligibility, giving rationale. | 6-7 |
| Information sources | 7 | Describe all information sources (e.g., databases with dates of coverage, contact with study authors to identify additional studies) in the search and date last searched. | Appendix pp5-6 |
| Search | 8 | Present full electronic search strategy for at least one database, including any limits used, such that it could be repeated. | Appendix pp5-6 |
| Study selection | 9 | State the process for selecting studies (i.e., screening, eligibility, included in systematic review, and, if applicable, included in the meta-analysis). | Appendix pp5-6 |
| Data collection process | 10 | Describe method of data extraction from reports (e.g., piloted forms, independently, in duplicate) and any processes for obtaining and confirming data from investigators. | 6-8 |
| Data items | 11 | List and define all variables for which data were sought (e.g., PICOS, funding sources) and any assumptions and simplifications made. | 6-8 |

| | | | |
|---|---|---|---|
| Risk of bias in individual studies | 12 | Describe methods used for assessing risk of bias of individual studies (including specification of whether this was done at the study or outcome level), and how this information is to be used in any data synthesis. | 8,9, data file |
| Summary measures | 13 | State the principal summary measures (e.g., risk ratio, difference in means). | 8 |
| Synthesis of results | 14 | Describe the methods of handling data and combining results of studies, if done, including measures of consistency (e.g., $I^2$) for each meta-analysis. | 7-8 |

Page 1 of 2

| Section/topic | # | Checklist item | Reported on page # |
|---|---|---|---|
| Risk of bias across studies | 15 | Specify any assessment of risk of bias that may affect the cumulative evidence (e.g., publication bias, selective reporting within studies). | 8,9, data file |
| Additional analyses | 16 | Describe methods of additional analyses (e.g., sensitivity or subgroup analyses, meta-regression), if done, indicating which were pre-specified. | 8 |
| **RESULTS** | | | |
| Study selection | 17 | Give numbers of studies screened, assessed for eligibility, and included in the review, with reasons for exclusions at each stage, ideally with a flow diagram. | 9,10,12, Fig 1 |
| Study characteristics | 18 | For each study, present characteristics for which data were extracted (e.g., study size, PICOS, follow-up period) and provide the citations. | Data File |
| Risk of bias within studies | 19 | Present data on risk of bias of each study and, if available, any outcome level assessment (see item 12). | Data file |
| Results of individual studies | 20 | For all outcomes considered (benefits or harms), present, for each study: (a) simple summary data for each intervention group (b) effect estimates and confidence intervals, ideally with a forest plot. | NA |
| Synthesis of results | 21 | Present results of each meta-analysis done, including confidence intervals and measures of consistency. | Table 1,2,3 |
| Risk of bias across studies | 22 | Present results of any assessment of risk of bias across studies (see Item 15). | 9,10, data file |
| Additional analysis | 23 | Give results of additional analyses, if done (e.g., sensitivity or subgroup analyses, meta-regression [see Item 16]). | NA |
| **DISCUSSION** | | | |
| Summary of evidence | 24 | Summarize the main findings including the strength of evidence for each main outcome; consider their relevance to key groups (e.g., healthcare providers, users, and policy makers). | 17-21 |
| Limitations | 25 | Discuss limitations at study and outcome level (e.g., risk of bias), and at review-level (e.g., incomplete retrieval of identified research, reporting bias). | 20 |
| Conclusions | 26 | Provide a general interpretation of the results in the context of other evidence, and implications for future research. | 20 |

| | | | |
|---|---|---|---|
| **FUNDING** | | | |
| Funding | 27 | Describe sources of funding for the systematic review and other support (e.g., supply of data); role of funders for the systematic review. | 20,21 |

*From:* Moher D, Liberati A, Tetzlaff J, Altman DG, The PRISMA Group (2009). Preferred Reporting Items for Systematic Reviews and Meta-Analyses: The PRISMA Statement. PLoS Med 6(7): e1000097. doi:10.1371/journal.pmed1000097

For more information, visit: **www.prisma-statement.org**.

### Box 1 - Search strategy for Review 1 of *P. vivax* Antimalarial Clinical Trials

**Search strategy**

All prospective *P. vivax* antimalarial clinical trials with a minimum of 28 days follow up, published between Jan 1, 1960 and Aug 23, 2021 were identified by the application of the key terms (listed below) through Medline (Pubmed), Web of Science, Embase and the Cochrane Central Database. Abstracts of all references containing any mention of antimalarial drugs were manually checked to confirm prospective clinical trials, with review of full text when needed.

To be eligible for inclusion in the current review, trials had to have been done since 1990, (after which adverse event detection and reporting was more standardised, include one or more treatment arm(s) in which patients were treated with either partial or fully supervised primaquine therapy, daily dosing for at least 5 days' duration. Primaquine administration had to commence within the first 7 days after starting blood schizontocidal therapy. Data from non-primaquine-containing arms in these studies were also extracted for comparative purposes, but restricted to patients treated with chloroquine, dihydroartemisinin-piperaquine or artemether-lumefantrine. Studies meeting the above criteria, but not reporting the presence or absence of adverse effects of treatment, were excluded.
The year of the study was taken as the year in which the paper was published, although the start and end date of patient enrolment were also recorded.
The review process was undertaken by two independent investigators who also performed data extraction (RJC and RNP), and is documented in more detail in Commons et al, Int J Parasitol Drug Drug Res 2017.

Previously registered at PROSPERO [CRD42016053228].

**Key terms:**

Literature search (conducted August 23, 2021) with the following key terms (version undertaken in Pubmed):

vivax AND (allopurinol OR amodiaquine OR atovaquone OR artemisinin OR arteether OR artesunate OR artemether OR artemotil OR atovaquone OR azithromycin OR artekin OR chloroquine OR chlorproguanil OR cycloguanil OR clindamycin OR coartem OR dapsone OR dihydroartemisinin OR duo-cotecxin OR doxycycline OR halofantrine OR lumefantrine OR lariam OR malarone OR mefloquine OR naphthoquine OR naphthoquinone OR pafuramidine OR piperaquine OR primaquine OR proguanil OR pyrimethamine OR pyronaridine OR proguanil OR quinidine OR quinine OR riamet OR sulphadoxine OR sulfamethoxazole OR tetracycline OR tafenoquine).

### Box 2 - Search strategy for Review 2 of Severe primaquine-associated Haemolysis

**A. Review strategy and search terms used**

**Search strategy**

All articles reporting at least one case of severe primaquine-associated haemolysis published between 1 January 1940 and 20 May 2020, were identified by the application of the key terms (listed below), through PubMed, Web of Science, Embase and the Cochrane Central Database.
Title and abstracts of all references were manually checked to confirm papers that reported data attributable to individual patients receiving primaquine for *P. vivax* radical cure or terminal prophylaxis. The review process was undertaken by six independent reviewers (DY, EG, KT, RJC, NMD, RNP), with discrepancies resolved by discussion.

The review was registered at PROSPERO [CRD42020196604].

**Key terms:**
Literature search (conducted May 2020) with the following key terms (version undertaken in Pubmed):

vivax and (hospital* or renal or dialysis or transfusion or severe or serious or haemolysis or hemolysis or fatal or death or died or methemoglobin* or methaemoglobin* Or 'cerebral complicat*' or convuls* or unconscious* or prostrat* or 'kidney injury' or 'renal failure' or 'renal impairment' or hemoglobinuria or haemoglobinuria or 'circulatory collapse' or shock or jaundice or hyperbilirubinemia or hyperbilirubinaemia or 'hepatic dysfunction' or 'liver dysfunction' or bleeding or hemorrhage or haemorrhage or thrombocytopenia or thrombocytopaenia or 'disseminated intravascular coagulation' or DIC or 'acute respiratory distress syndrome' or ARDS or 'pulmonary edema' or 'pulmonary oedema' or 'metabolic acidosis' or hyperlactat* or 'severe anemia' or 'severe anaemia' or hypoglycemia or hypoglycaemia or complicat*)

**B. Secondary searches:**

1. Review all articles included in the WWARN *P. vivax* clinical trial database from the first systematic review of serious adverse events

2. Identify additional studies, conference abstracts and unpublished works from reference lists of identified articles and documents

**Figure S1 - Total dose of PQ (mg/kg) administered (A) before first symptoms of haemolysis and (B) manifestation of severe haemolysis**

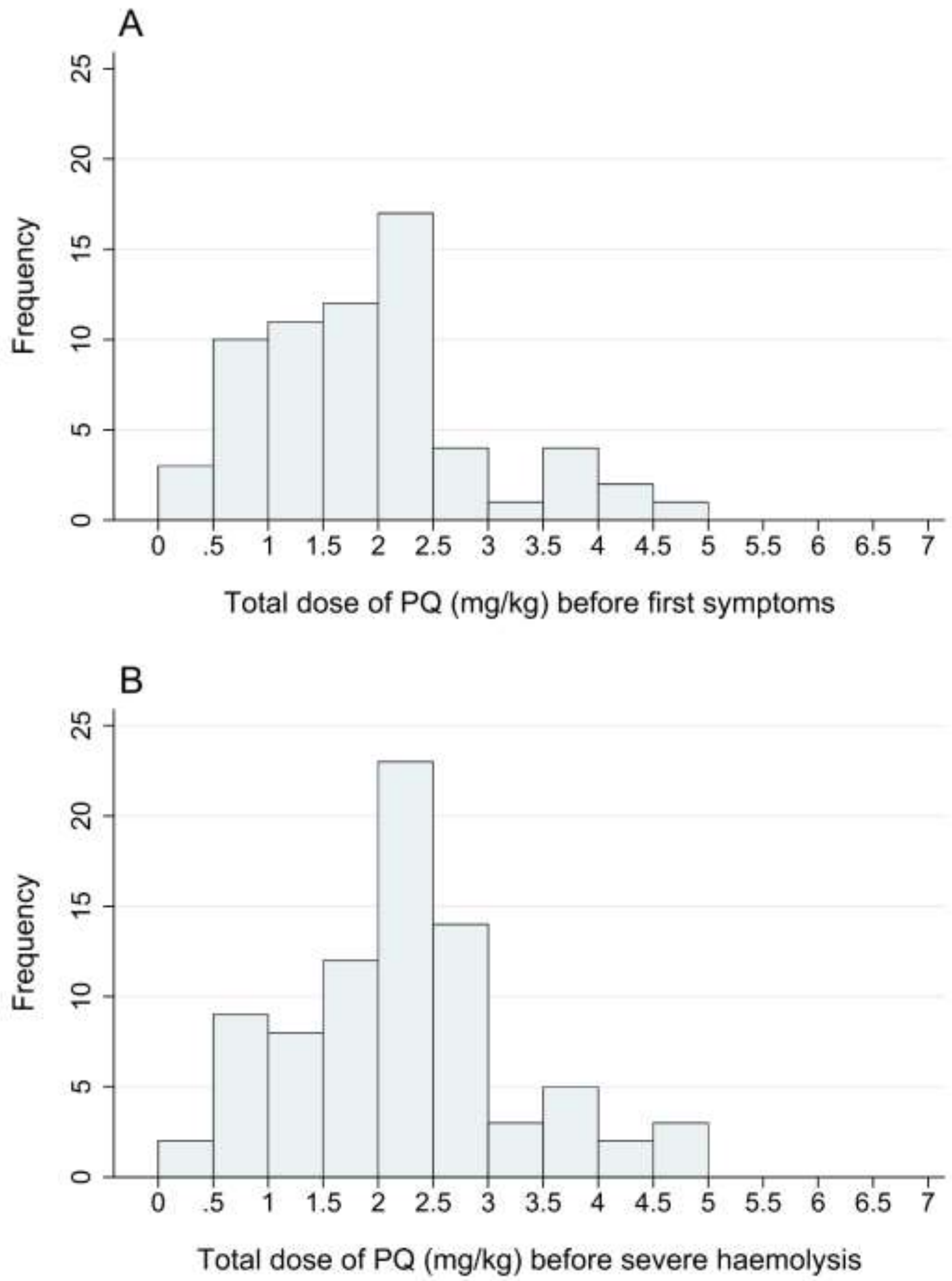

Footnote: Data restricted to the 101 cases with probable or possible severe PQ-associated haemolysis

### Example of Form for reporting drug-induced haemolysis following treatment of malaria

<table>
<tr><td colspan="2">REPORTER DETAILS</td></tr>
<tr><td>Name of reporter:____________________</td><td>Date of assessment: __ / __ / ___</td></tr>
<tr><td>Role / Function: _______________________</td><td rowspan="2">Report type:<br>☐ Initial ☐ Follow-up FU #: ____</td></tr>
<tr><td>Tel:_____________________________<br>Email:______________________________</td></tr>
<tr><td colspan="2">PATIENT DETAILS</td></tr>
<tr><td colspan="2">Patient initials: _______________ Medical ID number: __________________________<br>Sex: □ M □ F Age: _______ Date of Birth: __ /__ /___<br>Weight: _________ kg<br>Country: _____________________ Site: ________________________</td></tr>
<tr><td colspan="2">MALARIA DETAILS</td></tr>
<tr><td colspan="2">Treatment Indication: □ Acute malaria □ Terminal prophylaxis □ Other:___________<br>Date of Malaria Diagnosis: __ / __ / ___ Method: □ Microscopy □ RDT □ Other___<br>Malaria Species: □ Pf □ Pv □ Pm □ Po □ Pk □ Unknown<br>Baseline parasitaemia: _______________ per ul ++++ +++ ++ + □ Unknown</td></tr>
<tr><td colspan="2">TREATMENT DETAILS</td></tr>
<tr><td colspan="2">Schizontocidal Treatment: □ CQ □ AL □ DP □ Quinine □ None □ Other:____________<br>Hypnozoitocidal drug: □ PQ □ Tfq<br>Date Commenced: ___ /___ /____ Time Commenced: __ __ :__ __<br>Daily Dose of PQ or Tfq: ________.__ mg Calculated mg/kg dose:___________<br>Planned duration of PQ: ___________ days<br>Target total dose of PQ: ___.__ mg/kg<br>Number of doses taken before adverse event detected: _________<br>Did the patient take their PQ / TQ tablets with food? □ Yes □ No □ Unsure</td></tr>
</table>

| CONCOMITANT MEDICATION | | | | | |
|---|---|---|---|---|---|
| **Medication** | **Indication** | **Start date dd/mm/yyyy** | **Stop date dd/mm/yyyy** | **Dose** | **Frequency** |

### CLINICAL PRESENTATION

Date adverse event detected: __ / __ / __

Temperature:____.___ C Pulse: _____ per min Resp rate: ____ per min

| **Symptom** | **Severity (see criteria table)** | **If present, date of onset** |
|---|---|---|
| Abdominal pain | None 1 2 3 4 | ___/___/20___ |
| Nausea | None 1 2 3 4 | ___/___/20___ |
| Unable to eat | None 1 2 3 4 | ___/___/20___ |
| Vomiting | None 1 2 3 4 | ___/___/20___ |
| Back pain | None 1 2 3 4 | ___/___/20___ |
| Breathlessness | None 1 2 3 4 | ___/___/20___ |
| Dizziness | None 1 2 3 4 | ___/___/20___ |
| Fatigue | None 1 2 3 4 | ___/___/20___ |
| Severe Conjunctival Pallor | Yes / No | ___/___/20___ |
| Fever | Yes / No | ___/___/20___ |
| Jaundice | Yes / No | ___/___/20___ |
| Tachycardia | Yes / No | ___/___/20___ |
| Cyanosis | Yes / No | ___/___/20___ |
| Dark (red or black) urine | Colour:______<br>(See Hillman chart) | ___/___/20___ |
| Other: ________________ | None 1 2 3 4 | ___/___/20___ |

### NARRATIVE

_____________________________________________________________________________

_____________________________________________________________________________

_____________________________________________________________________________

_____________________________________________________________________________

_____________________________________________________________________________

_____________________________________________________________________________

### RELEVANT MEDICAL HISTORY

| Medical condition | Start date dd/mmm/yyyy | Stop date dd/mmm/yyyy | Ongoing |
|---|---|---|---|
| | | | ☐ |
| | | | ☐ |
| | | | ☐ |
| | | | ☐ |

### RELEVANT INVESTIGATIONS

| Haemoglobin | Date | Result |
|---|---|---|
| Pre-treatment | __ / __ / 202__ | Hb: __ . __ g/dL |
| At time of event | __ / __ / 202__ | Hb: __ . __ g/dL |
| Current | __ / __ / 202__ | Hb: __ . __ g/dL |
| Nadir | __ / __ / 202__ | Hb: __ . __ g/dL |

**Max fall in Hb:** Hb: __ . __ g/dL

**Max fractional fall in Hb** ____.__ %

#### G6PD STATUS

**G6PD test**: □ Quantitative □ Qualitative □ Unknown □ Not Done

**Name of test**: ___________________________

**Date of testing**: __ / __ / __ **Time of testing**: __ __:__ __

**Quantitative result**: _____ . __ U/g Hb □ Deficient □ Intermediate □ Normal

**Qualitative result**: □ Deficient □ Normal □ Indeterminant

**Genotyping**: □ NA □ Normal □ Variant _______________

#### OTHER LABORATORY TESTS

| Test | Pre-treatment | At time of event | Follow-up | Not Available |
|---|---|---|---|---|
| | __ / __ / ___ | __ / __ / ___ | __ / __ / ___ | |
| **WBC:** | __________ x$10^9$ | __________ x$10^9$ | __________ x$10^9$ | □ |
| **Plt:** | __________ | __________ | __________ | □ |
| **Na:** | _______ µmol/L | _______ µmol/L | _______ µmol/L | □ |
| **K:** | _______ µmol/L | _______ µmol/L | _______ µmol/L | □ |
| **Urea:** | _______ µmol/L | _______ µmol/L | _______ µmol/L | □ |
| **Total Bili:** | _______ µmol/L | _______ µmol/L | _______ µmol/L | □ |
| **Unconj Bili:** | _______ µmol/L | _______ µmol/L | _______ µmol/L | □ |
| **ALP** | _______ µmol/L | _______ µmol/L | _______ µmol/L | □ |
| **ALT** | _______ µmol/L | _______ µmol/L | _______ µmol/L | □ |
| **LDH:** | _______ µmol/L | _______ µmol/L | _______ µmol/L | □ |
| **Met Hb:** | ____.____ % | ____.____ % | ____.____ % | □ |

**Other Investigations:**

### ADVERSE EVENT CLASSIFICATION

| | | |
|---|---|---|
| **Severity**<br>Maximum Graded Symptom | ☐ Grade 1 ☐ Grade 2 ☐ Grade 3 ☐ Grade 4 | |
| **AESI**<br>**Haemolysis**<br>□ Yes<br>□ No | Any of the following:<br>□ Grade 3 or 4: fatigue, dizziness, breathlessness (onset after starting PQ)<br>□ Severe pallor or jaundice<br>□ Dark urine: Hillman >7<br>□ Fall in Hb > 3 g/dL<br>□ Hb <7g/dl | |
| **SAE**<br>Meets “serious” criteria<br>□ Yes<br>□ No | □ Death | |
| | □ Life threatening | |
| | □ Hospitalisation or prolongation of hospitalisation | Admission date: __ / ___ / 202__<br>Discharge date: __ / ___ / 202__ |
| | □ Persistent or significant disability | |
| | □ Is a congenital abnormality / birth defect | |
| | □ Is an important and significant medical event | |
| **Relationship (causality) to PQ** | ☐ Not related ☐ Unlikely related<br>☐ Possibly related ☐ Probably related ☐ Definitely related | |

### CLINICAL MANAGEMENT

| | | |
|---|---|---|
| **IV Fluids** | □ Yes □ No | |
| **Blood transfusion** | □ Yes □ No | Number of units: _____ Date: __/__/202_ |
| **Dialysis** | □ No | □ Peritoneal dialysis □ Haemodialysis |
| **Changes to PQ** | □ No change □ Withhold □ Cease □ Restart | |
| | If Restarted: Date Restarted: __/__/202_<br>□ Same dose □ Modified dose<br>Dose: ___.__ mg □ 1x Day □ 2x day Duration: ___ Days | |

### NARRATIVE:

______________________________________________________________________________

______________________________________________________________________________

| |
|---|
| ______________________________________________________________________________<br>______________________________________________________________________________<br>______________________________________________________________________________<br>______________________________________________________________________________ |
| **OUTCOME** |
| □ Recovered / Resolved |
| □ Recovering / Resolving |
| □ Not recovered / Not resolved |
| □ Recovered / Resolved with sequelae Specify:___________________________ |
| □ Fatal: Date of death: ___ / ___ / 202__<br>Cause of death: ________________________________ |
| □ Unknown |
| **CLINICIAN RESPONSIBLE FOR THE REVIEW** |
| **Name :** Date: __ / __ / 202__ |
| **Role :** |
| **Address :** |
| **Mobile:** _______________________________ |
| **Email :** _______________________________ |
| **Signature:** ___________________________________________ |
